# Ribosome heterogeneity arising from common and rare rRNA sequence variants affects diverse human phenotypes

**DOI:** 10.1101/2025.09.02.25334953

**Authors:** Daphna Rothschild, Anil Raj, Jordan Brown, Nathaniel Thayer, Manuel Hotz, David Hendrickson, Jonathan K. Pritchard, Maria Barna

## Abstract

rRNA genes exhibit intra-individual hyper-variability and an outstanding question is their role in human health and disease. These include variants positioned at enigmatic regions of rRNA named Expansion-Segments (ESs) that protrude from the core of the ribosome, with poorly understood functions. In this study, we analyze rRNA variants in the UK Biobank population, revealing that common rRNA variations that give rise to *ribosome subtypes*, affect human physiology and rare *rRNA-mutations* affect diseases. We developed a Ribosome-Variation-Analysis (RiboVAn) method, identifying *ribosome subtypes* as heritable and a larger proportion of low-heritability *rRNA-mutations*. The heritable variants included ones in es15l associated with adiposity, es39l with body dimensions, and es27l with blood-related traits and diseases. Variant-chromosome specificity is observed where *ribosome subtypes* are linked to distinct acrocentric chromosomes. Burden analysis linked *rRNA-mutations* to diverse diseases including cancer and acute myocardial infarction. These findings causally link rRNA variation to human traits, disease, and establish that ESs have distinct and important functions in human physiology.

## Introduction

Ribosomal RNA (rRNA) genes present significant challenges for genetic studies due to its highly repetitive nature, with hundreds of copies arranged in tandem arrays on the five acrocentric chromosomes ^1^. Moreover, since rRNA is the most abundant RNA molecule in every cell, reaching 90% of total RNA ^2^, rRNAs are routinely depleted and discarded from standard transcriptome sequencing analyses. Population genetic studies have also largely ignored rDNA regions, as they are absent from Single Nucleotide Polymorphisms (SNP) arrays given their repetitive nature. However, rRNA genes are high copy number paralogs which raises the possibility that sequence variations between copies may form new types of heterogeneous ribosomes that have specialized functions during translation ^3^. This idea was suggested already in the 1970s, where it was reported that there are variations between the rDNA repeats ^4^.

Recent studies, including our previous work, analyzed the population of the 1,000 Genomes Project ^5^ and reported that humans carry hundreds of rDNA sequence variants ^6–8^. Yet prior studies relied on short read sequencing which is depleted of highly GC-rich rRNA genes ^9^, and studies reported discordant findings ^6,7^. In our previous study, using newly available long-read sequencing in the 1,000 Genomes Project together with developing an accurate variant calling algorithm for paralog genes, Reference Gap Alignment (RGA), we curated the rRNA sequence variants found in the 1,000 genome project ^8^. We found that the majority of rRNA variants are in the form of insertion deletion variants, including numerous new variants that were missed by short-reads. We identified tens of highly abundant variants, found in many rDNA copies in the genome, which we termed ribosome subtypes, and over a thousand low abundant variants found only in a few copies spread across the rDNA arrays.

These ribosome subtypes may be new types of heterogenous ribosomes having distinct functional roles in normal physiology whereas low abundant variants could be envisioned as acquired mutations that may have the potential to be associated with specific pathological features. Moreover, we showed that the variants were found in translating ribosomes by successfully sequencing full length rRNA (Ribo-RT) from translationally active polysome fractions in both human embryonic stem cells (hESCs) and K562 cells ^8^. We curated the variants into an atlas of rRNA sequence variations found in translating ribosomes. Using long-read DMS-seq in the hESC, we showed that the ribosome subtypes had different ribosome structures which could potentially lead to changes in ribosome function. To assess if ribosome subtypes could have a role in physiology we tested if ribosome subtypes expression levels are dynamic and by analyzing the GTEx dataset ^10^, we showed that ribosome subtypes had differential expression across tissues belonging to different developmental germ layers ^8^. In addition to normal physiology, we hypothesised that rRNA-mutations found normally in low abundance may potentially affect disease and by analyzing the TCGA cancer dataset ^11^ we found specific low abundant variants that became elevated in cancer biopsies ^8^.

Interestingly, tens of highly abundant rRNA variants found in translating ribosomes were found in expansion segment (ES) regions of the 28S. ESs are enigmatic hyper-variable insertions within the rRNA, located on the outer surface of the ribosome, that vary in length and between eukaryotic species ^12^. In humans, expansion segments (ESs) have nearly doubled the rRNA sequence length compared to prokaryotic rRNAs, with several ESs extending to hundreds of nucleotides. Since ESs are not evolutionary conserved, this has led to two notions: either they lack critical functions because they are outside of the conserved ribosome core or alternatively that they have more regulatory functions on the ribosomes that may even be species specific ^12^. Yet it remains difficult to study ES functions since rRNA genes can not be genomically engineered given hundreds of copy numbers in the genome. Moreover, studying ESs is challenging because they are flexible and their structures remain unresolved by cryo-EM. Still, our previous work using DMS-seq showed that sequence variants at ESs change their structures around sites of variations. While very little is known about the function of ESs, multiple recent studies suggest they are important^13,14,15,16,17^. One of the longest ESs, named es27l which is about 700 bases long, was shown to bind MetAP and to be important for translation fidelity^13,14,15^. Moreover, it was shown that mRNAs bind ESs^16^, possibly regulating their translation, including Hoxa9 mRNA which was shown to bind specifically es9s^17^. Given that highly abundant variants were found in ES regions it is possible that these variants alter the function of ES regions and control ribosome function.

While overall changes in rRNA, largely at the level of copy number variation, were associated with human health and disease ^18–21^, it remains an outstanding question whether ESs have functional roles and whether common variants found within them affect human phenotypes. Moreover, rare rRNA mutations could also possibly contribute to disease. While editing the rRNA is not possible given their high copy numbers, the UK Biobank (UKBB), with its extensive whole-genome sequencing (WGS) data ^22^, provides an unprecedented opportunity to dissect germline from somatic mutations and test the causal role of rRNA variations on human traits. In Genome Wide Association Studies (GWAS), germline mutations in protein coding sequences establish causal links between genes to phenotypes whereas mutations in non-coding regions are interpreted as associations. Here we study if rRNA variants affect human traits by studying if variants are germline or somatic of origin and focus on atlas variants found and confirmed in translating ribosomes ^8^. Indeed, recent work leveraging WGS has shown that germline variations in rDNA affect complex human traits^19,23,24^. Here, we investigate whether distinct common ribosome subtypes confer specialized functions that contribute to ribosome heterogeneity in normal human physiology, and study rRNA-mutations effects on diseases.

Using the WGS from the UKBB and our curated atlas rRNA variants we show that ribosome subtypes have different effects on human traits mediated by functional changes in ES regions. Moreover, we find that rRNA-mutations are associated with a wide-range of diseases. These findings not only demonstrate the critical importance in ES function for human physiology but also define how variations in the most highly paralogous genes in the human genome are associated with numerous human traits.

## Results

### A Pipeline for Associating rRNA Variants with Human Traits

The human rDNA loci are organized in hundreds of tandem repeats on the short arms of five acrocentric chromosomes (**Figure 1A**). In our previous study, we generated a comprehensive atlas of rRNA sequence variants that are confirmed to be present in translating ribosomes ^8^. Here, we investigated the role of rRNA variants in human health and by focusing our analysis on variants from our atlas, we ensure that we are studying variations found in translating ribosomes (**Figure 1A**). Here, we move beyond the associations found in our previous work to ask if these variants affect human traits. Specifically, since the direction of information flows from DNA to phenotype, establishing that germline mutations in rRNA associate with a phenotype implies their causal effect on the tested phenotype ^25,26^ (**Figure 1B**).

**Figure 1:**
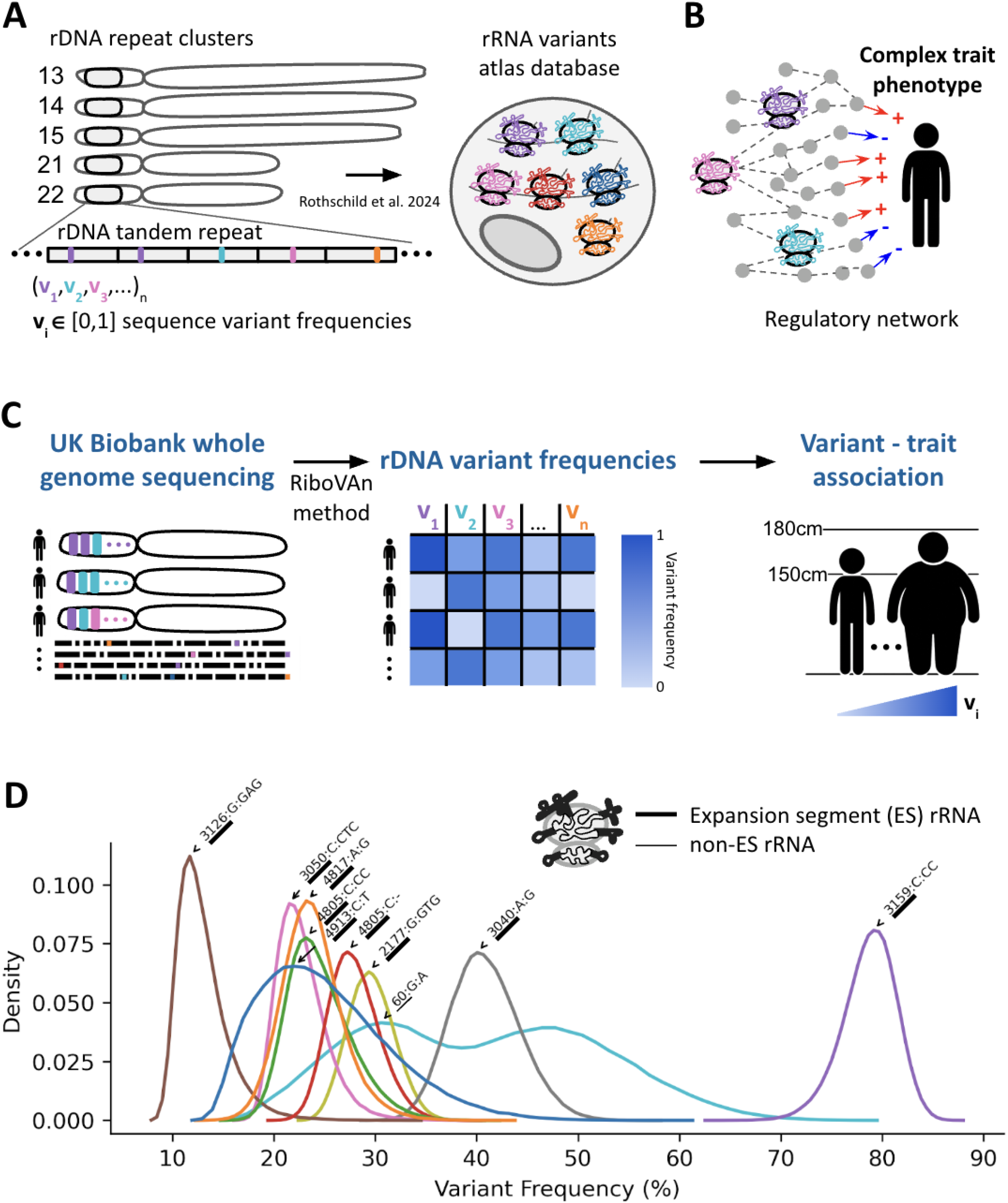
An rRNA sequence variants analysis pipeline. (A) Structure of the rDNA acrocentric chromosomes (13, 14, 15, 21, and 22) depicting the repeat clusters. The rRNA variants atlas database is shown, illustrating the sequence variant frequencies identified in translating ribosomes, as previously reported by Rothschild et al. (2024) ^8^.(B) Conceptual framework illustrating the hypothesis that rRNA variants affect complex trait phenotypes through regulatory networks. The figure highlights the potential connections between rRNA variants and various human health traits. (C) Workflow for the analysis conducted in this study. Whole genome sequencing data from the UK Biobank is mapped to the rRNA variant atlas using the RiboVAn method for variant calling. The resulting rDNA variant frequencies are then associated with human traits. (D) Distribution of variant frequencies for the top ten rRNA variants identified in the study. The density plot illustrates the frequency of variants, with annotations indicating the location of rRNA variants on the ribosome, emphasizing that most variants are located within expansion segments (ES).

Since rRNA genes are highly repetitive, short read sequencing cannot identify the chromosomal location of variants. Instead, we use short-reads to call variant frequencies within the rRNA. For calculating variant frequencies in short-read data, it is necessary for short-reads to align to longer sequences than the short-reads themselves. Since, UKBB short reads are 150 bases long and we expanded the published rRNA atlas ^8^ with 150 bases of non-variant reference sequences on both 5’ and 3’ ends in order to map reads to the atlas (**Data S1, Methods**). Yet in order to find single nucleotide variants after mapping to variant combinations there needs to be additional conversion from variant combinations to single nucleotide variants. Here we developed the Ribosome Variant Analysis (RiboVAn) pipeline to calculate atlas single nucleotide variant frequencies from short-read data.

RiboVAn reports the fraction of variant frequencies observed per individual for every atlas variant (**Figure 1C**). We created lookup tables, one for each ES/non-ES region, which contain for every combination of variants the positions and single nucleotide variants that are found in combination. With these tables, after mapping reads to the ES resolution atlas which contain combinations of variants, we sum nucleotide variants using the mapped reads and the conversion tables to report nucleotide relative frequencies. We released the RiboVAn tool as part of this study (**Methods** and **Data Availability**).

With this method we calculated the nucleotide variant frequencies for 928 variants, 318 variants from the 18S and 610 variants from the 28S, in 487,822 individuals in the UKBB cohort. Importantly, given GC-biases found in short-read data, as we later describe here, we analyze sequencing variant read depth statistics which we use to study the noise of variant calling. The resulting variant frequency table from RiboVAn is then used for further heritability analysis and association with phenotypic traits.

To control for technical biases, particularly the under-sequencing of GC-rich regions by Illumina platforms, we compared variant frequencies from UKBB short-reads to those from long-read sequencing of the hESCs from our atlas. We observed that highly abundant variants found in hESCs were all highly abundant variants in the UKBB and in agreeing frequencies, but we observed much lower agreement for the hESC low abundant variants (**Figure S1A**). When analyzing the UKBB rRNA variant sequencing depth, we found that discordant variants, those found in low frequency in the hESC and high frequency in the UKBB had low sequencing read count and likely resulted from high noise (**Figure S1B**).

Since hESCs had similar variant frequencies to the average sample in the UKBB, we next asked if variant frequencies vary within the population. Highly-abundant variants showed higher variation in the population and the top 10 most variable ones all belonged to the 28S large subunit rRNA gene (**Figure 1D**). These variants also had low noise levels when examining their sequencing raw read count. Surprisingly, we observed a wide distribution of variant frequencies in the UKBB population in these top variants. For example, the variant G:A at position 60 of the 28S varied dramatically in frequency between individuals from 14% to 80% of the copies carrying the G to A variation (**Figure 1D**, cyan histogram showing the distribution of 60:G:A variant). This makes it particularly interesting to ask how such variability in the population affects human phenotypes. Moreover, in agreement to our previous study, most of these variants were located in the 28S rRNA ESs (**Figure 1D**, ES/non-ES are annotated on the cartoon ribosome). Indeed, these included high-frequency variants identified in our previous work that form distinct structural subtypes within the ES regions es15l, es27l and es39l. We conclude that highly abundant variants mostly within ES regions display high variations between the UKBB cohort.

### rRNA Variants Include Both Heritable and Somatic Variations

Next we asked if rRNA variants are heritable, germline variations. To distinguish between germline and somatic variants, we assessed heritability using 177 monozygotic twins within the UKBB. We found that a subset of rRNA variants is highly heritable (**Table S1**). This included 53 variants with high correlation between twins (Spearman R>0.8), confirming their germline origin (**Figure 2A**, an example of one highly heritable rRNA variant, **Figure B2**, dashed line indicating Spearman correlation exceeding 0.8 between twins). This finding is in agreement with Rodriguez-Algarra et al. reporting germline rRNA variations ^24^. However, unlike the previous study, we observed that the overall distribution of twin-variant-correlations (which we use to estimate heritability) is skewed towards lower values, suggesting that a majority of rRNA variants are likely somatic (**Figure 2B, Figure S2A** an example of one lowly heritable rRNA variant).

**Figure 2:**
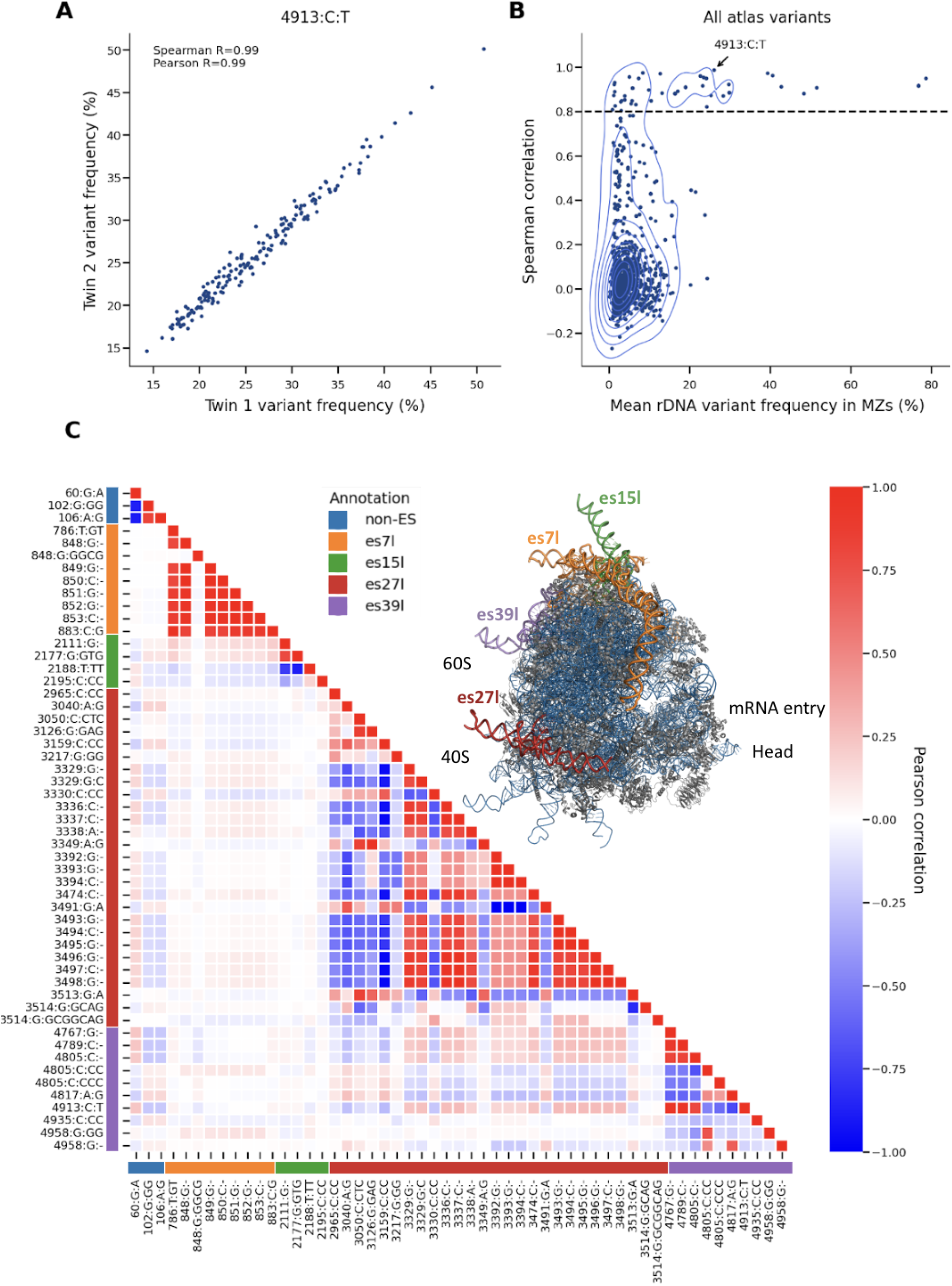
rRNA variants include germline and somatic sequence variations. (A) Scatter plot showing the correlation of variant frequencies between monozygotic twins for the variant 4913:C:T of the 28S gene. The Spearman correlation coefficient (R = 0.99) and Pearson correlation coefficient are indicated, demonstrating high heritability for this specific variant. (B) Contour plot illustrating the Spearman correlation of all atlas variants against their mean rDNA variant frequency in monozygotic twins. The dashed line indicates the threshold for significant heritability, with the variant 4913:C:T from panel (A) highlighted. (C) Heatmap displaying high heritable variants (from panel B) correlation to one another. The color gradient represents the Pearson correlation values. A structure of a ribosome (PDB 4v6x ^27^) with ES/non-ES region indicating the specific expansion segments (es7l, es15l, es27l, es39l) and non-ES variants in different colors. These colored annotations or ribosome regions are also annotated in the rows and columns of the heatmap.

We asked if low-heritability was driven by noise. Indeed, variants belonging to regions with lower sequencing depth showed low-heritability, but this did not account for the entire signal (**Figure S2**). Specifically, we compared heritability to the variant read-depth in the twins. Out of 928 rRNA variants tested, 354 variants had low read-depth (<5000 reads in average mapped to the rRNA variant, **Figure S2** showing variant read depth). Using all variants, we found moderate correlation between heritability and sequencing-depth (Pearson R=0.25 between the mean variant read depth and twin-variant-correlation). After removing low read-depth variants, the remaining 570 variants showed relatively low correlation between heritability and sequencing depth (Pearson R=0.12 between the mean variant read depth and twin-variant-correlation). This supports that overall low heritability remains after controlling for GC biases, which likely reflects somatic mutations. We conclude that both heritable and somatic rRNA variants exist.

rDNA copy numbers were recently shown to be heritable and associated with different traits, mostly blood count traits ^23^. Accordingly, we asked if rDNA copy numbers are associated with sequence variations. Heritable highly abundant ribosome subtype variants showed low correlation with rDNA copy number variations while the low frequency rRNA-mutations showed varying correlations with rDNA copy numbers (**Figure S4**). This result indicates that changes in rDNA copy numbers and ribosome subtype frequencies are largely independent. However, given high correlations between copy numbers and some rRNA-mutations, this result suggests in the process of rDNA copy number changes somatic mutations may occur at specific rDNA hotspots.

The most highly heritable variants are predominantly located in the 28S rRNA, specifically within four out of twenty nine mammalian ESs: es7l, es15l, es27l and es39l (**Figure 2C**, see ribosome structure video). These variants include mostly highly-abundant variants, termed as ribosome subtypes, that show large abundance-variability within the cohort (**Figure 1D**). We next focused on these 53 most highly heritable variants (Spearman R>0.8), as their association with human traits imply a causal link between rRNA sequence variations and human physiology, thereby establishing biological functions for ESs.

In agreement with our previous study, these variants form haplotypes, combination of co-inherited variants, within the ES region but have much lower linkage to other regions outside individual ES regions (**Figure 2C**, ES region annotations marked in different colors and with a ribosome cartoon). This suggests that ESs are largely independent from one another and could potentially have different functions. Additionally, we observed some weak linkage in between regions es27l, es39l and position 60 of the 28S and almost no linkage to es7l and es15l (**Figure S3**). In accordance with our previous study, while es7l is the longest ES, its variants are lowly linked to other ESs. Together, this supports that ES regions could have independent functions with potential cooperations between some ESs.

### Heritable Expansion Segment Variants Associate with Distinct Human Traits

To test if rRNA variants affect human traits, we performed an association study between the common heritable variants and a wide range of human traits, including anthropometric traits, blood counts and diseases in the UKBB (**Table S2, S3, Methods**). Strikingly we observed many significant rRNA-variant-trait associations.

Blood calcium levels for example, showed no significant associations to any of the heritable rRNA variants (**Figure 3A**). In contrast, we found significant associations between ES variants and key physiological traits (**Figure 3B, 3C**, regression analysis P-value<0.05 after Bonferroni correction for the number of independent traits times the number of variants see **Methods**). Surprisingly, different variants belonging to different ESs were associated with the traits. Variants in es27l are associated significantly with IGF-1 levels, a crucial growth hormone involved in development and adult tissue maintenance ^28^. In contrast, es15l and es39l variants are associated with standing height.

**Figure 3:**
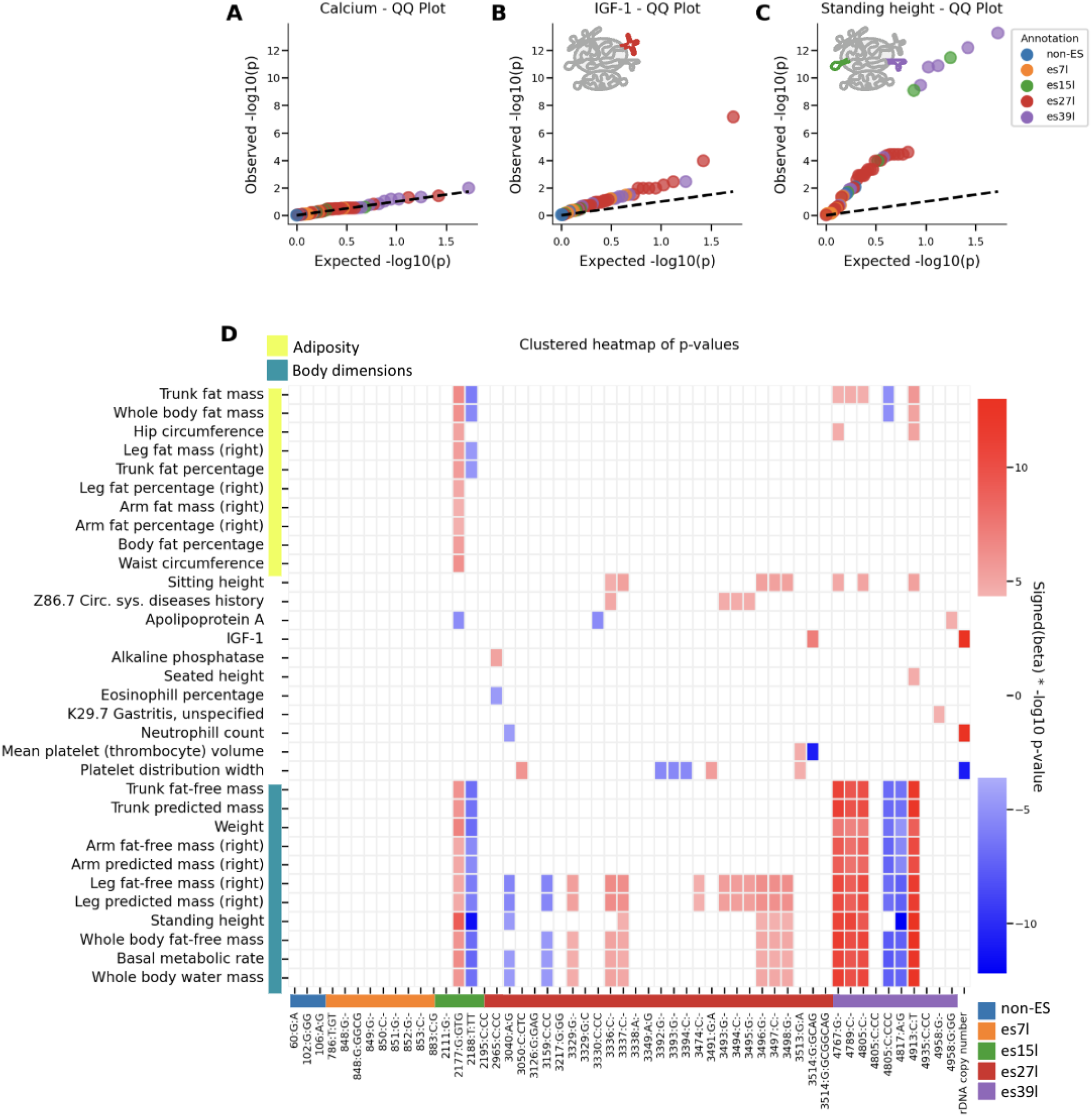
Expansion segments are associated with human traits. (A) Quantile-quantile (QQ) plot for the association of rRNA variants with blood calcium levels. The plot indicates no significant association, as evidenced by the distribution of observed versus expected p-values which follows the dashed line of random chance p-values. (B) QQ plot showing the association of rRNA variants with IGF1 levels. The plot highlights significant associations, with expansion segments annotated in different colors corresponding to their respective ES. es27l is annotated in red on the ribosome cartoon. (C) Same as (B) for standing height. es15l and es39l regions are annotated in green and purple on the ribosome cartoon. (D) Clustered heatmap of p-values for the association of rRNA variants and copy numbers with various traits related to adiposity and body dimensions. For visualization purposes, minus log10 p-values were capped at 13 (**Table S2, S3** include the full regression results). Rows indicate traits and columns belong to variants. The rows are clustered and are annotated in yellow or teal if they belong to adiposity or body dimension related traits. Except for the rDNA copy number column, columns are annotated in color by their ES location. The heatmap visually represents the strength of associations, with color gradients indicating significance levels for each trait. The colors red and blue indicate the sign of the linear regression beta where red indicates positive association with the trait and blue for negative association.

Importantly, GWAS that aim to identify variants in the genome that affect human traits, mostly identify variants in non-coding parts of the genome ^29^. These are usually interpreted as variants that are found in genomic proximity to the genes that have a causal effect on the focal trait of interest. However, unless these variants are located in known enhancer or promoter regions, they remain hard to interpret. In contrast, the tested atlas variants are coding variants, found in translating ribosomes as opposed to a genomic regulatory region. This means that germline variations in rRNA affect human traits. Furthermore, given that the linkage between ESs is relatively low (**Figure 2C**) this shows that different ESs have divergent effects on traits.

When taking a broader view of trait associations to rRNA variants, we observed clear clustering by trait category (**Figure 3D**, traits are hierarchically clustered by and annotated in yellow and teal). Variants in es15l were most strongly linked to traits related to adiposity, while variants in es39l showed the strongest association with body dimension traits, including standing height. Interestingly we also observed different blood related traits that were associated with es27l, including platelet distribution width, and two disease associations; history of circulatory diseases to es27l and gastritis to es39l. This grouping highlights that es15l, es27l, and es39l perform unique functions. Interestingly, while two ESs, es15l and es39l are associated with some overlapping traits, es27l had strong association to other traits and es7l did not associate significantly to any trait. The overlap in es15l and es39l traits stem from es15l associating with both body dimension and adiposity related traits. Overall this suggests that es15l may have a broader ribosomal function in human health compared to more nuanced but important ribosome functions of es27l and es39l which affect distinct aspects of human health and disease.

Notably, our regression models included rDNA copy number estimates and in agreement with previous reports we found that rDNA copy numbers were associated with different blood traits (**Figure S5**). Yet, we observed little overlap between the traits associated with the sequence variants, and those associated with rDNA copy numbers (**Figure 3D**). This shows that observed variant-trait associations are largely independent of rDNA copy number variations.

Having observed that heritable ribosome subtypes are associated primarily with normal physiological traits, we next asked whether ribosomal heterogeneity arising from low-frequency mutations found in a few rDNA copies is associated with disease. Mutation burden analysis is frequently used to increase the power of testing for association between low frequency mutations in protein coding genes with complex traits. Typically, in such an analysis, different mutations in the same gene are grouped together and we test for an association between the number of mutations within the gene and the trait. To this end, instead of testing for association between hundreds of low frequency mutations in rDNA with diseases, we accumulated many low frequency mutations within defined rDNA regions, one for each ES and non-ES region of the rRNA, and asked if mutation burden at these regions associates with diseases (**Figure S6, Table S4** for ES, non-ES annotations, **Methods**).

For each rDNA region, we tested if this region carries high mutation frequency and calculated the Major Allele (MAF) per rDNA region, which is the most common genetic variant in the population. Consistent with our previous analysis here, es7l, es15l, es27l, es39l, and pre-es5l which includes position 60 of the 28s had a low MAF representing hyper variability at these regions (see **Figure 2B,C**). Additionally, we observed that instead, for many rDNA regions, there is very low sequence variation, with one major allele being prevalent in the population and with a major allele close to 100% (**Figure S6**). For these constrained regions, any deviation from 100% tests the burden of mutations associated with disease incidence. This analysis revealed that mutations in rDNA regions from both the 18S and 28S with low mutation incidence are associated with increased risk for multiple diseases (**Figure 4**). These surprisingly included common and rare diseases such as type 2 diabetes, acute myocardial infarction, congenital birth disorders, and cancers (**Figure 4**). Similar to our previous variant-level analyses, we controlled for the effect of rDNA copy numbers on disease, and found that for common UKBB diseases and clinical biomarkers there was also association with rDNA copy numbers but not for rare UKBB diseases. Of note, since the mutation incidents at these regions is low, we added stringent coverage cutoffs to ensure that the coverage does not suffer from low sequencing read depth (**Methods**). It is important to note that the UKBB is a general population cohort, not a disease-focused one; therefore, the disease associations we identified likely represent an underestimation of the impact of rRNA variations on human disease. We conclude that rRNA variations as a whole display striking association with many human traits.

**Figure 4:**
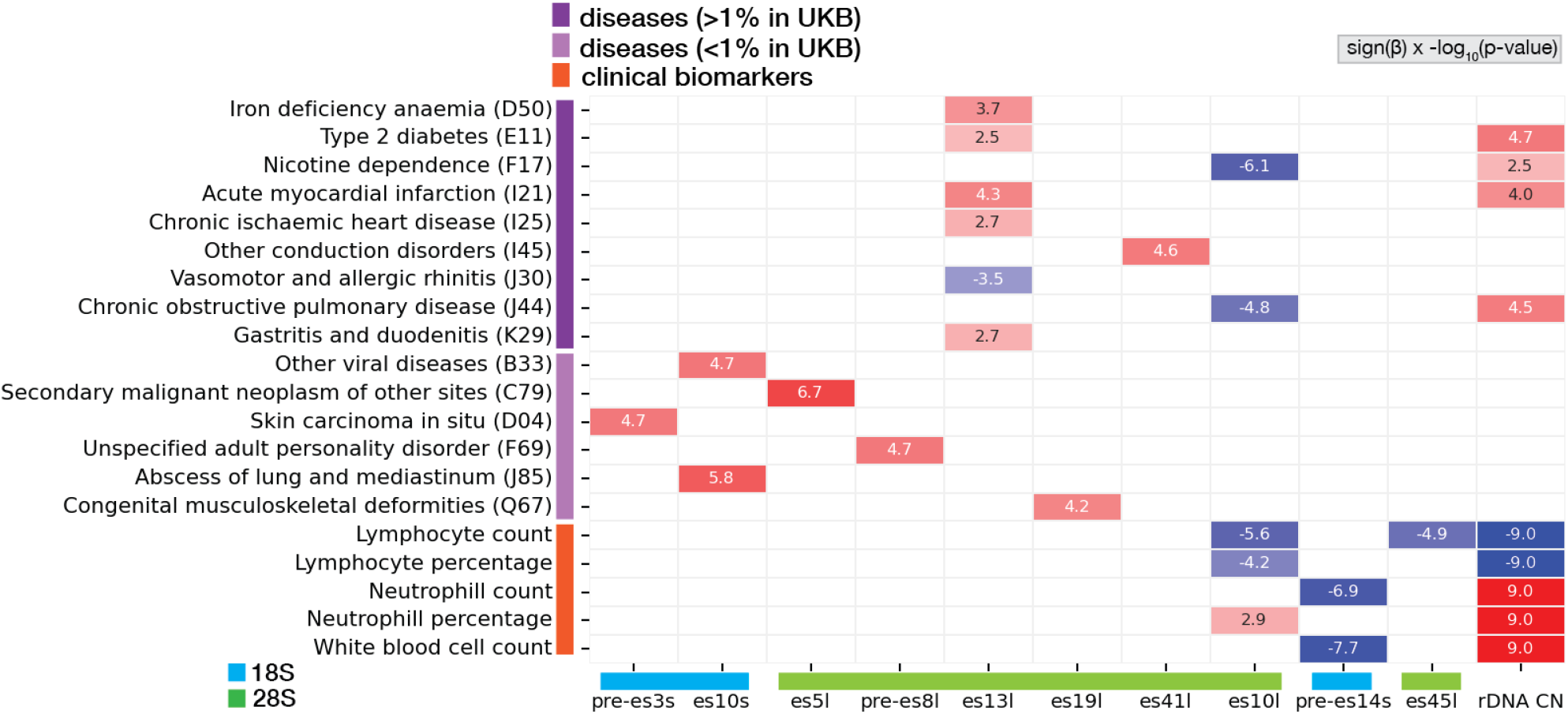
Rare variant burden in ES and non-ES regions and complex traits. Heatmap of significant associations between the MAF of the rDNA region with diseases and clinical biomarkers. The last column of the heatmap shows significant associations of rDNA copy number (CN) with the same traits. Numbers in each cell are sign(**β**) x −log10(p-value) of the association.

### es15l and es39l Variants Exhibit Distinct Linkage Patterns to Acrocentric Chromosomes

In our previous study we found that variants were clustered in arrays and some were chromosome specific ^8^. Here given that the UKBB has short-read data, we cannot map variants to their genomic location. Instead, we ran a GWAS analysis to associate rRNA variant frequencies to the rest of the genome. This allowed us to investigate the genomic origins of these heritable variants by analyzing their linkage to the five acrocentric chromosomes (13, 14, 15, 21, and 22).

As expected, all variants associated with the acrocentric chromosomes and only with the acrocentric chromosomes (**Figure 5**). This further strengthened our causality results as we do not find other genetic associations with the ribosome subtypes outside the acrocentric chromosomes harboring the rDNA loci. When focusing on variants belonging to different ESs, we observed different linkage patterns. We focused on the top trait associated variants that belonged to different ESs, 2177:G:GTG from es15l, 3514:G:GCAG from es27l, and 4913:C:T from es39l. Interestingly, variants from es15l and es39l had a stronger association to one chromosome: The es15l variant showed the strongest linkage to chromosome 21 (**Figure 5A**) and the es39l variant was most associated with chromosome 14 (**Figure 5C**). In contrast, the es27l variant was strongly linked to multiple chromosomes 14, 15, 21, and 22 (**Figure 5B**). Given that es15l and es39l variants showed higher chromosome specificity, this could potentially point to regulation of adiposity and body dimension traits by regulating chromosome specific rRNA arrays.

**Figure 5:**
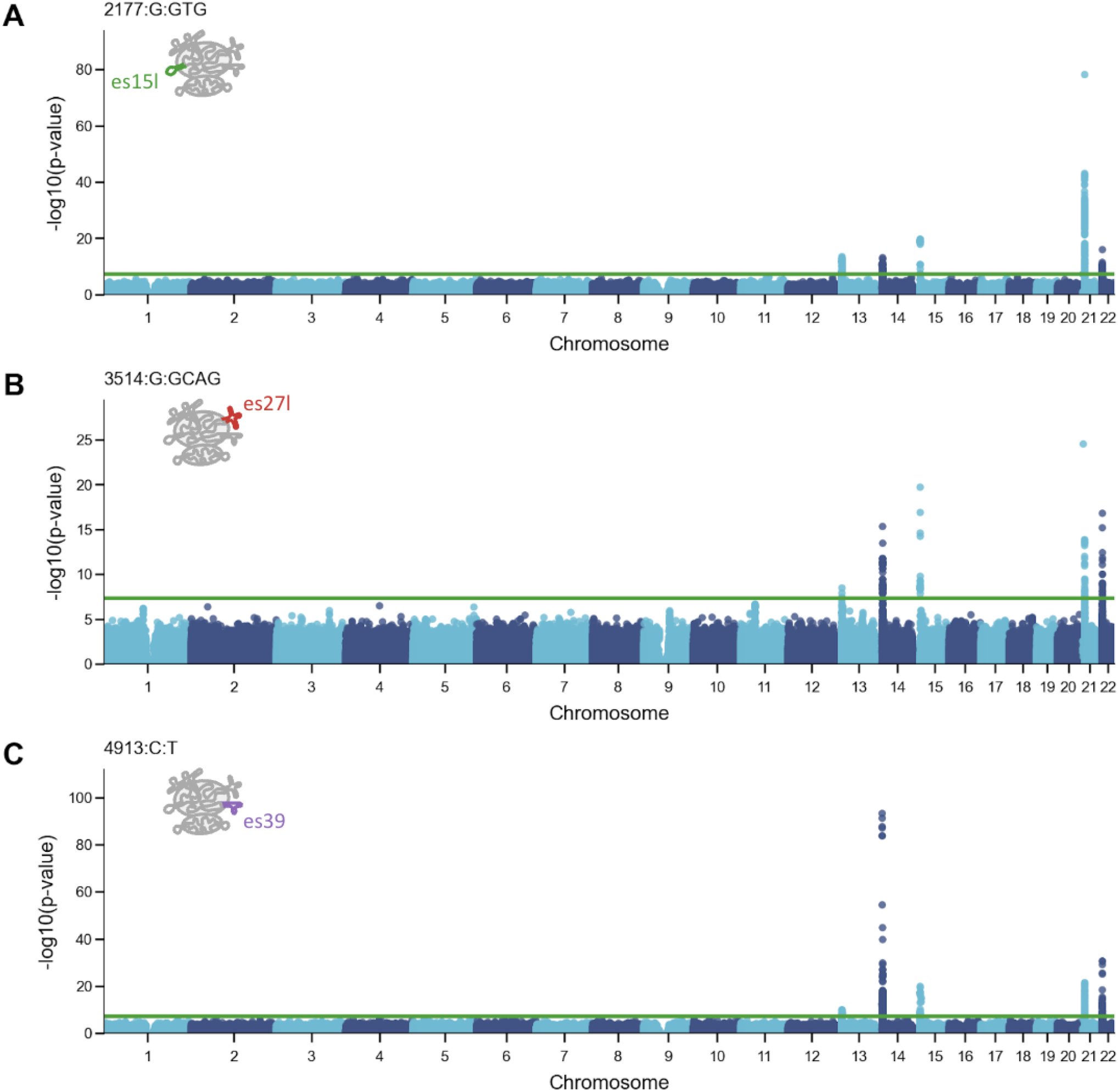
Varying genomic linkage between rRNA variants and acrocentric chromosomes. (A) Manhattan plot displaying the association of the top variant located in es15l (2177:G:GTG) with specific chromosomes. The y-axis represents −log10(p-value) for the association. es15l is annotated in green on the ribosome cartoon. (B) Similar to (A) for the top variant located in es27l (3514:G:GCAG). es27l is annotated in red on the ribosome cartoon. (C) Similar to (A) for the top variant located in es39l (4913:C:T). es39l is annotated in purple on the ribosome cartoon.

Variant-chromosome specificity is interesting given the high propensity of translocations in acrocentric chromosomes that would homogenize the rRNA array across chromosomes. Specifically, Robertsonian translocations are the most common translocations in humans, happening in about 1 in every 1,000 newborns ^30^. These translocations occur between the short arms of two acrocentric chromosomes at specific sites ^31^. Here our data indicates that variant-chromosome specificity still occurs in the background of translocation events. However, since the UKBB has short-read data, we could not test if chromosome variants are clustered in rRNA arrays. Together, our data supports chromosome specificity of some variants while other variants are more evenly distributed across multiple chromosomes, consistent with the high tendency of translocation events in these chromosomes.

## Discussion

This study establishes that rRNA variations are associated with human traits and diseases. Furthermore, rRNA variants located in ESs have clear phenotypic implications. While the precise mechanisms remain to be elucidated, our findings demonstrate that es15l influences both adiposity and body dimensions, es39l primarily affects body dimensions traits, and es27l impacts blood phenotypes and disease susceptibility (**Figure 3**). ESs may expand our knowledge of how the ribosome itself can be linked to metabolic traits. The outstanding question is how different ESs function. The observed associations between the variations in the ES to different phenotype categories, i.e. body dimensions, adiposity and blood traits, raises the hypothesis that ESs show specificity as well as cooperativity between ESs. It may be that different ESs facilitate interactions with specific proteins, mRNAs, or even subcellular organelles that control translation. Moreover, ESs have also been shown to directly interact with each other, for example by direct tertiary RNA interactions ^43^ potentially working in concert for regulating important aspects of translation control linked to human physiology.

There is accumulating literature that supports a critical role for ribosome function in hematopoiesis and that mutations in ribosome components in ribosomopathies lead to defects in hematopoiesis, including bone marrow failure ^23,32–37^. These include Shwachman-Diamond syndrome affecting neutrophils, caused by a mutated ribosome biogenesis factor ^36^, Diamond Blackfan Anemia caused by mutations in ∼20 ribosomal proteins that causes selective defects in erythroid progenitors ^38^, specific thrombocytopenia caused by mutation in SLFN14 that causes defects in rRNA regulation which affects platelets ^33^. rRNA levels are also associated with hematopoiesis where hematopoietic transcription factors were shown to regulate rRNA transcription ^34^ and at the genome level we and others have found that copy number variations in rRNA affect hematological profiles (**Figure S5**) ^23^. Together this supports the notion that tight regulation of ribosome function has broad roles in hematopoiesis, and future studies are needed to shed light on how common variations in es27l and rRNA-mutations affect hematopoiesis in particular.

Additionally, we found variants in es15l and es39l that affect body dimension related traits, adiposity as well as linking these variants to specific chromosomes (**Figure 3**,**5**). This raises multiple interesting directions. In particular, in different ribosomopathies, patients have short stature ^37,39,40^ and it would be interesting to study if these diseases carry rRNA mutations that control body size. Moreover some variants in es15l and es39l are linked to specific chromosomes. A new study found that rRNA gene arrays can be silenced and reactivated by DNA methylation and de-methylation, which can be inherited ^41^. It may be that there is chromosome level regulation of rRNA variant transcription that is needed for metabolism or growth. There is a growing appreciation for tight metabolic control that is governed at the translation level ^42^.

This study found that rare rRNA-mutations are associated with a variety of diseases, including cancer (**Figure 4**). This adds to the growing work that associated rRNA variants and copy numbers with cancer ^8,20,21,44–47^. In our previous work we found cancer specific rRNA mutations that were normally lowly expressed in control samples but were elevated in cancer biopsies in 11 different cancer types ^8^. Elevated ribosome biogenesis is a hallmark for cancer, and future investigations in dedicated cancer cohorts will be necessary for uncovering if elevated expression of particular cancer associated rRNA-mutations are driven by rDNA expansion in copy numbers coupled with rRNA-mutations or if TFs control rRNA transcription of a subset of rDNA copies with cancer associated rRNA-mutations.

It is not well understood whether increases in rDNA copy numbers result in the production of more ribosomes. Our results do not provide an answer to this question, but interestingly the set of phenotypes found associated with rDNA copy number changes and rRNA variant frequency changes are mostly non-overlapping. This supports a high degree of uncoupling between rRNA variations and rDNA copy number changes. It is interesting to study the drivers of rRNA-mutations. The highly accessible chromatin state of active rDNA loci might render them more susceptible to DNA breaks and subsequent erroneous repair, contributing to the accumulation of somatic variants over time. This damage may accumulate with age and it would be interesting to further study rRNA-mutations in aging. Moreover, in our previous work, we identified that the vast majority of variants are in the form of short insertion deletions (indels) ^8^. With the rRNA being enriched with GC rich homopolymer tracts, indels may be a result of sequences prone to replication slippage, which could explain the high prevalence of specific indels. In contrast, some of the rRNA-mutations had a modest correlation with rDNA copy numbers which supports studying if there are rDNA hotspots for somatic mutations that are selected for growth and cell fitness.

Together, using human genetics our findings highlight that ribosome heterogeneity at the level of rRNA is meaningful for human physiology and disease. Moreover, as ESs have had elusive functions on the ribosome, our studies highlight the functions of specific ESs that affect human traits ranging from body height, adiposity, and blood phenotypes. This opens new directions for their study by pinpointing which physiological processes are guided by these unique rRNA regions associated with an expansion in ribosome size across evolution.

## Supporting information

Data S1 - containing a fasta file of extended ES resolution rRNA atlas - with 150 bases of the reference sequence flanking 5' and 3' ends

## Data and code availability

In this publication we analyzed the whole genome sequencing and phenotypes of the UKBB cohort and the whole genome sequencing of the H7-hESC cell line. Both UKBB and H7-hESC are published datasets. The H7-hESC rDNA is available under BioProject ID PRJNA926787 under accession numbers SRR23196516 (H7-hESC whole genome sequencing). We generated an extended ES atlas with extended 150bp reference sequences as Extended Data to this publication (**Data S1**). This is the same ES atlas as we previously published ^8^ with 150bp flanking reference sequences to allow longer 150bp reads found in the UKBB dataset to map to the atlas. The RiboVAn pipeline is available at https://github.com/daphnar/rRNA/RiboVan

## Methods

### Datasets used in this study

The datasets analyzed in this study are the WGS and the genotype data available for 487,822 samples in the UKBB, and rDNA single nucleotide variant frequencies from the rRNA atlas, found in table **Data S1** ^8^.

### Mapping UKBB WGS short-reads to the rRNA atlas

In order to obtain rRNA variant frequencies, we first converted the raw CRAM file for each UKBB participant to fastq reads mapping to rRNA, using the following samtools ^48^ command:

~~~
>> samtools view -C -o “${sample}.ribo” --reference “$reference”
-M -L “$ribo_bed” -@ 20 “${cram_file}”
>> samtools fastq -c 9 -0 /dev/null --reference “$reference”
“${sample}.ribo” -o “${sample}.ribo.fq.gz”
~~~

Here `{sample}` indicates the participant ID, `{cram_file}` is the input CRAM file and `{reference}` is the hg38 human genome reference fasta file.

For the ribosome coordinated, i.e. $ribo_bed, we used the following:

chr21:8205987-8219245

chr21:8250196-8255929

chr21:8389034-8402286

chr21:8433221 8446515

chr22_KI270733v1_random:122272-135588

chr22_KI270733v1_random:167354-179715

chrUn_GL000220v:1105423-118723

chrUn_GL000220v:1149395-161745

Next, we map these rDNA reads to an atlas of sequence variation within annotated expansion segments (ES) and non-expansion segments (non-ES) in the 18S and 28S subunits of the 45S rDNA unit. The atlas annotates the 18S subunit into 9 ES and 10 non-ES segments, and the 28S subunit into 19 ES and 19 non-ES segments. Since the UKBB WGS short-reads are 150 bases long, we used an ES atlas with extended 150bp reference sequences that we generated as part of this paper, **Data S1**. To generate it we concatenated on the 5’ and 3’ end the rRNA reference sequence flanking the ES atlas ^8^.

### Ribosome Analysis Variation (RiboVAn) pipeline

To calculate rRNA variant frequencies for every sample, we performed the following steps:

1. For every ES/non-ES region as defined in Table S4 and in accordance with our previous study ^8^, we found all nucleotide variations associated with the ES/non-ES variant. We stored this information as a lookup table that mapped an ES variant to a list of all nucleotide variants found in this ES variant.
2. Using Bowtie2 ^49^ mapper, we mapped short reads to the ES resolution atlas with flanking extended 150bp reference sequences (**Data S1**).
3. We counted reads with perfect alignment to the ES resolution atlas variants.
4. Then we used the lookup table from step (1) and converted the ES resolution variant counts to the nucleotide resolution variant counts.
5. We divided the nucleotide variant counts by the total number of reads mapped to the ES/non-ES region to get the relative abundances of nucleotide variants.

The RiboVAn pipeline is available at https://github.com/daphnar/rRNA/RiboVan

### Rare variation burden within 18S and 28S rDNA subunits

First, we retrieved all alignments of the rDNA reads to the atlas using the command `bwa mem -a`. Then, for each ES / non-ES segment, we resolved the read alignments into counts for its annotated alleles. Since the following algorithm is applied independently for each segment, we drop indexing by segment for notational convenience. Let *A* be the set of alleles in the atlas for segment *s* and *R*_*p*_ be the set of reads from participant *p* mapping to segment *s*. We defined perfect read-allele alignments as those with no mismatches or gaps in the cigar strings of the alignments, when restricting to the coordinates of the segment *s* and read positions with base quality scores greater than 20.

For each read *r* ∈ *R*_*p*_, we defined an alignment vector α_*r*_ ∈ {0, 1}^|*A*|^, where α_*ar*_ = 1, if the read *r* perfectly aligned to allele *a*. Some reads did not perfectly align to any allele in *A*. Given our stringent base quality filters, we hypothesized that novel variants (rather than sequencing errors) predominantly contributed to this set of reads. However, we ignored these reads for our analyses due to the difficulty of accurately discovering variation in highly GC-rich regions using short-read sequencing data. Some reads perfectly aligned to more than one allele and were, thus, denoted as the set of *multi-mappers*. We ignored multi-mapper reads in order to be conservative in our estimates of segment-level variation.

The remaining reads perfectly aligned to exactly one allele in *A*; 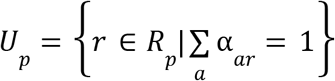. We denoted *U*_*p*_ as the set of *unique mappers* and computed 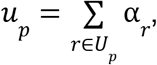 the count vector of unique mappers for participant *p* at segment *s*. We observed that some alleles had little *unique mapper* support in the entire UKBB cohort.

We defined 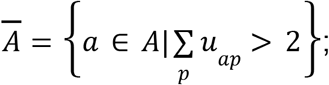 the set of alleles that had at least 2 *unique mappers* across the entire cohort. Since short-read data in this cohort cannot distinguish between the remaining alleles, *A* − *Ā*, we retained one allele from this remaining set, the most abundant allele as previously reported in our atlas,and excluded the rest. Thus, the set of resolvable alleles for this segment, *Ā* ^+^= *Ā* + *the retained most abundant allele*. This operation changed the dimensions of the alignment vector for each read; 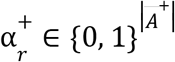. Our set of unique mappers now includes reads that, previously, perfectly aligned to multiple alleles that were deemed indistinguishable; 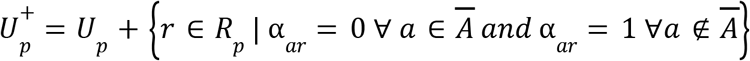. We updated our count vector of unique mappers as 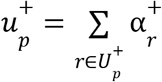.

We computed the read depth for the segment *s* in participant *p* as 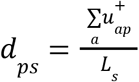, where *L*_*s*_ is the length of the segment. We excluded segments with a median read depth less than 30, since variant calls in these segments would be less accurate. For the remaining segments, we estimated the frequency of allele *a* in participant *p* as 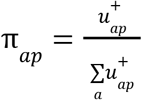. We estimated the copy number of each allele as 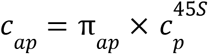, where 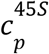 is the estimated 45S copy number for participant *p*. We set allele copy numbers less than one to zero, 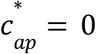 *if c*_*ap*_ < 1, acknowledging that these alleles are likely somatic variation in low-frequency clones in participant *p*. Our final estimate of the frequency of allele *a* in participant *p* was defined as 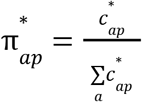.

### Genetically related participants in UK Biobank

The UKBB provides genetic relatedness information for 107,162 pairs of participants, covering a total of 147,753 participants. Participant pairs with kinship > 0.4 were considered monozygotic twins. Participant pairs with kinship between 0.17 and 0.35, and birth years differing by more than 18 years were considered parent-offspring pairs. We used these criteria to identify 177 monozygotic twins and 1012 trios with WGS data in the UK Biobank cohort.

### Regression analysis with rRNA variant frequencies

We tested for an association between rRNA variants and quantitative traits using an ordinary least square linear regression model, and for an association with disease incidence using a logistic regression model (using python’s `statsmodels.api` package). We restricted our regression analyses to 402,610 unrelated White British individuals to control for population structure, and filtered based on sample QC characteristics as previously described in Sinnott-Armstrong et al., 2021^50^.

The UKBB includes hundreds of ICD10 codes coding for diseases, but the UKBB is a general population cohort and is not a targeted disease cohort. In order to minimize tests, we focused on ICD10 codes with at least 30,000 cases (6% prevalence). Moreover, to correct for multiple hypotheses, we applied Bonferroni correction using the number of rRNA variants tested times the number of diseases.

For logistic regressions, rRNA variants and covariates were normalized to one (by subtracting the mean and dividing by the standard deviation). The statsmodels.api.Logit models for some variant-traits pairs failed to converge; we excluded these tests in our analyses.

In the regression models we controlled for age, sex, age -sex interaction, age^2^, rDNA copy number, population structure and batches. Specifically, we estimated rDNA copy number by the fraction of reads mapped to the rDNA out of total reads, as described previously ^23^. In addition to restricting our analyses to unrelated White British individuals, we add the 20 first genetic principal components to the model to account for population structure. Categorical batches for WGS measurements were added to the model using one hot encoding.

### GWAS analysis

GWAS analyses were performed on genotyped data using the autosomal bi-allelic SNPs as described in Mostafavi et al., 2020 ^51^. For associating variant frequencies with the rest of the genome, we used plink2 ^52^ with the following bash command for every chromosome-variant pair. The variant_id file stores variant frequencies for a selected rRNA variant and the rest of the chromosome files contain genotyped data.

~~~
phenofile=“${variant_id}.txt”
pgenfile=“chr${chromosome}.pgen”
psamfile=“chr${chromosome}.psam”
pvarfile=“chr${chromosome}.pvar”
famfile=“chr${chromosome}.fam”
bimfile=“chr${chromosome}.bim”
covarfile=“ukbb_20pc.txt”
plink2 --pgen “$pgenfile” --bim “$bimfile” --fam “$famfile”
--pheno “$phenofile” --covar “$covarfile” --glm hide-covar
--covar-variance-standardize --out “$outfile” --threads 16 --vif 999
~~~

## Acknowledgments

We thank Yuxiang Chen from the Barna lab for creating the video and caption of the ribosome structure. We thank the Barna and Pritchard group members for discussions. M.B. is supported by the National Institutes of Health grant R01HD086634. J.K.P. and D.R are supported by RO1 HG014005 and HG008140.

## Author contributions

D.R. conceived and directed the project, designed and conducted all computational analyses, interpreted the results, and wrote the manuscript. A.R, J.B, N.T, M.H, and D.H designed and conducted the rare-variant burden analysis, interpreted the results, and wrote the manuscript. M.B. and J.K.P. conceived and directed the project and analyses, designed the analyses, interpreted the results, and wrote the manuscript.

## Declaration of interests

A.R, J.B, N.T, M.H, and D.H were employees of Calico Life Sciences LLC at the time of this study. All authors declared no competing interests.

**Figure S1:**
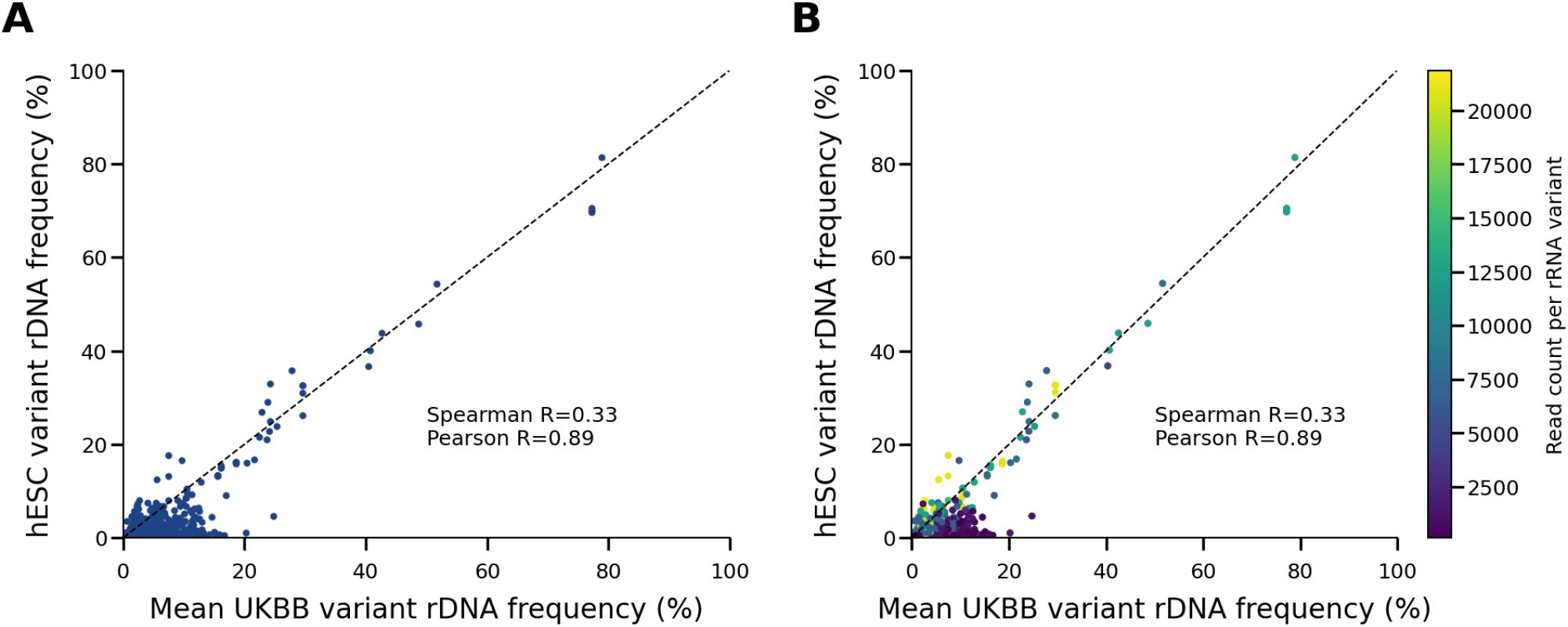
rRNA variants frequencies in hESC cell line and average UKBB profile agree for high coverage sequencing variants. (A) Scatter plot comparing the genomic rRNA variant profile of two datasets: mean UKBB and the hESC cell-line. Spearman and Pearson correlations are presented. (B) Similar to (A) but variant frequencies are colored by the number of reads that were mapped to the rRNA variant. A color bar indicating read count is presented.

**Figure S2:**
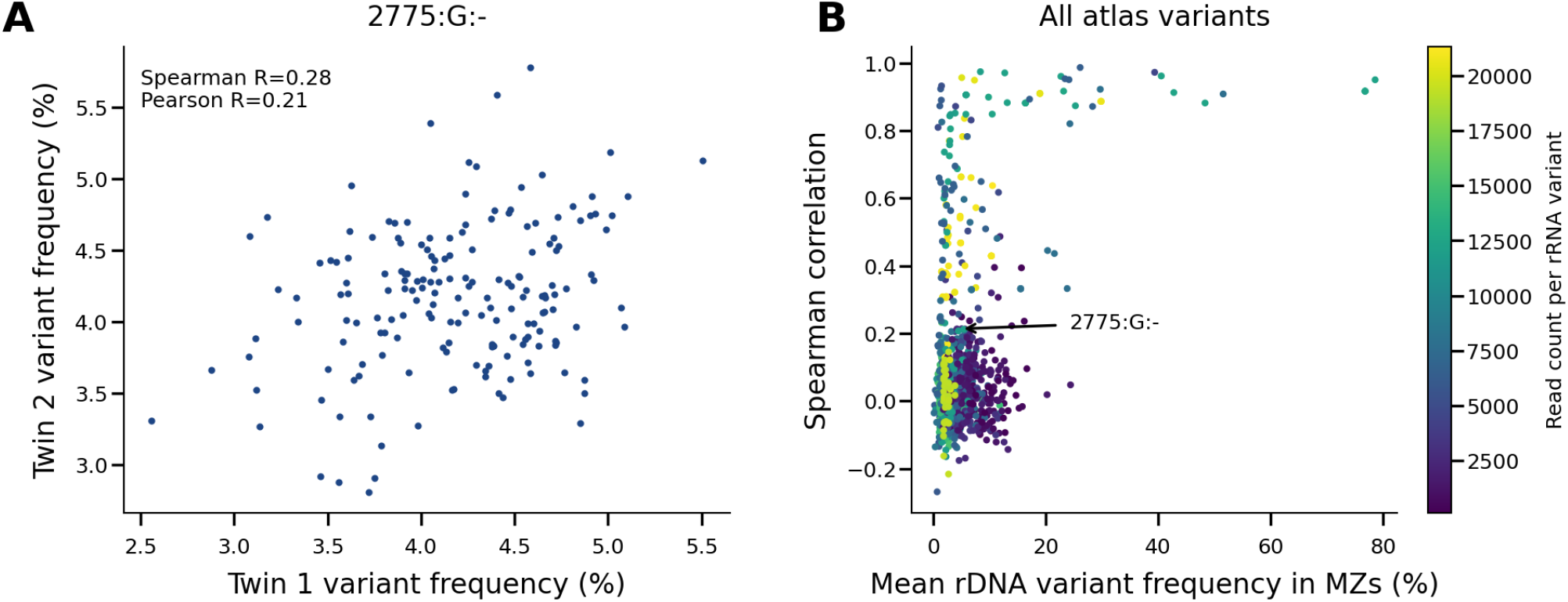
rRNA variants contain somatic sequence variations. (A) Scatter plot showing the correlation of variant frequencies between monozygotic twins for the variant 2775:C:T of the 28S gene. The Spearman correlation coefficient (R = 0.28) and Pearson correlation coefficient are indicated, demonstrating low heritability for this specific variant. (B) Scatter plot showing the Spearman correlations of all atlas variants against their mean rDNA variant frequency in monozygotic twins. The Spearman correlation for variant frequencies are colored by the number of reads that were mapped to the rRNA variant. A color bar indicating the mean read count per sample is presented. The variant 2775:C:T from panel (A) highlighted.

**Figure S3:**
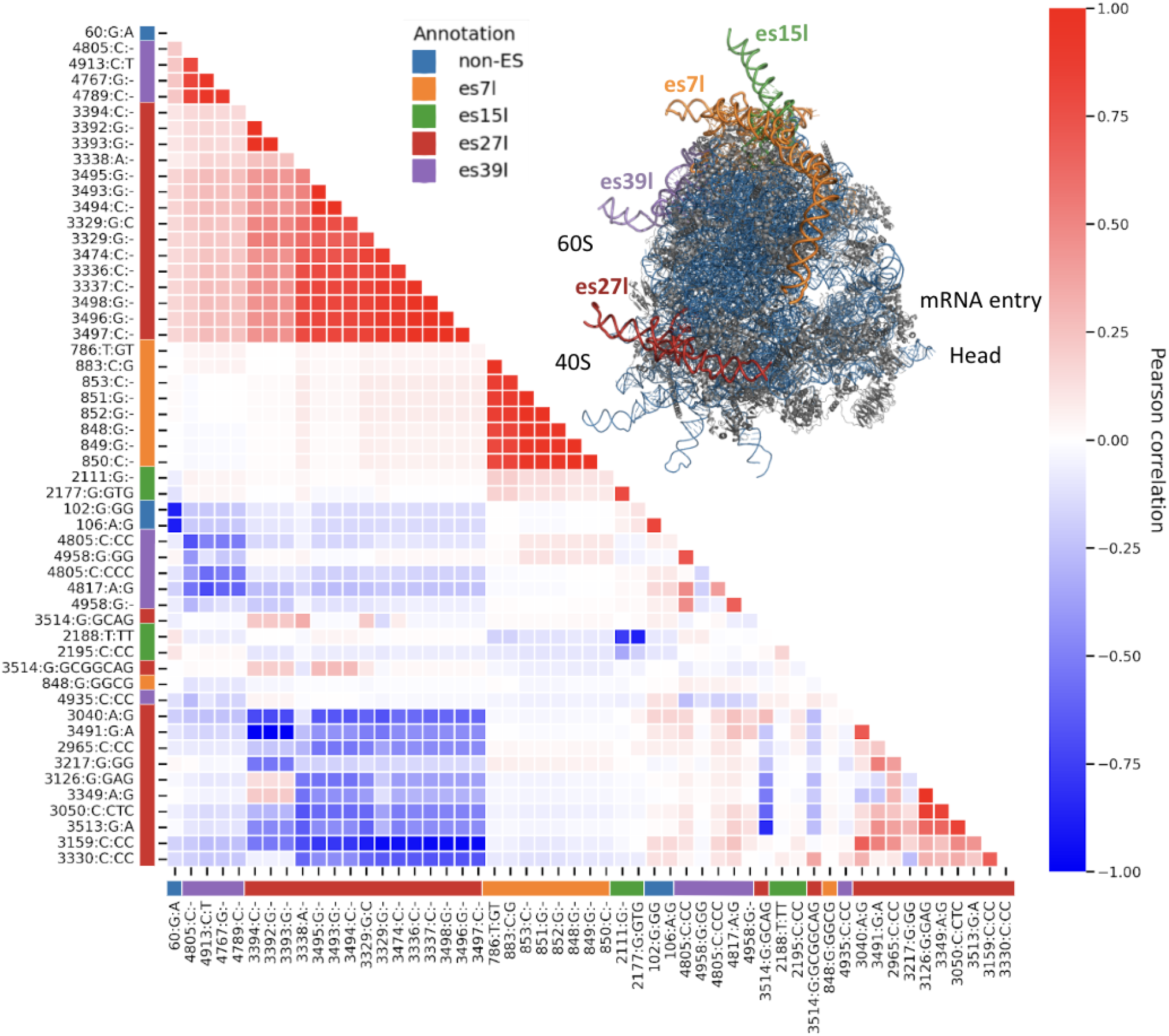
rRNA variants shows some haplotype linkage between rRNA regions. Heatmap displaying high heritable variants correlation to one another after hierarchical clustering. The color gradient represents the Pearson correlation values. A structure of a ribosome (PDB 4v6x ^27^) with ES/non-ES region indicating the specific expansion segments (es7l, es15l, es27l, es39l) and non-ES variants in different colors. These colored annotations or ribosome regions are also annotated in the rows and columns of the heatmap.

**Figure S4:**
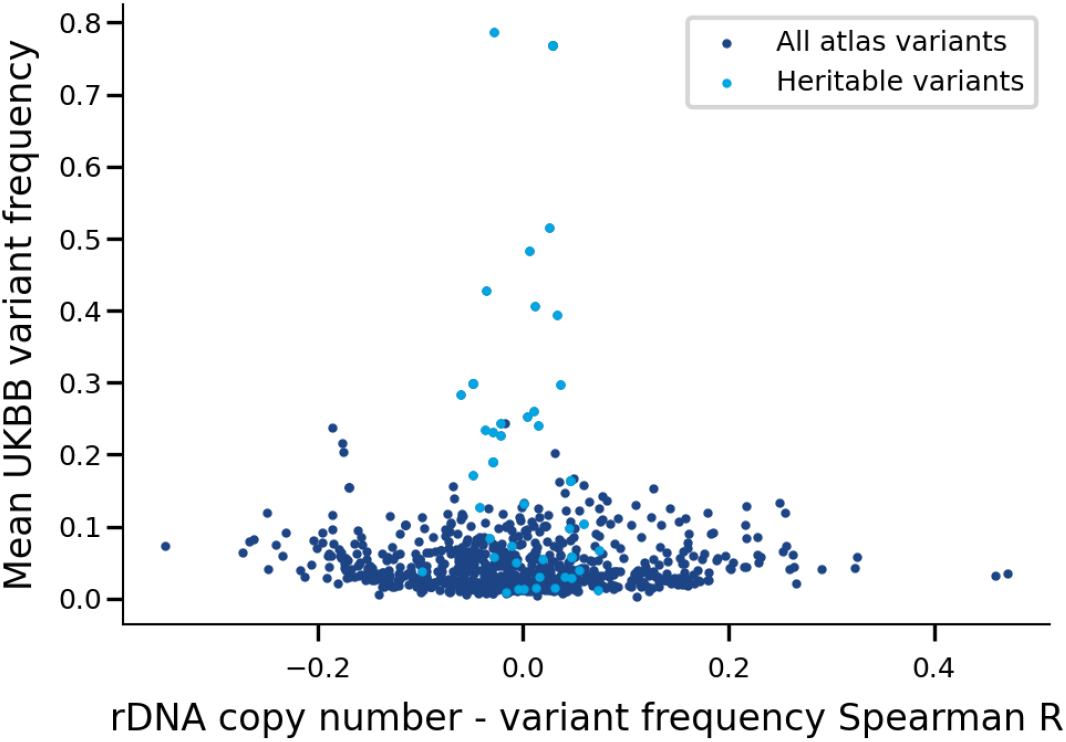
Heritable rRNA variants and rDNA copy numbers are not well correlated. Scatter plot displaying the Spearman correlation between rDNA copy number estimates and rRNA variant frequencies on the X-axis, and the mean UKBB rRNA variant frequency on the Y-axis.

**Figure S5:**
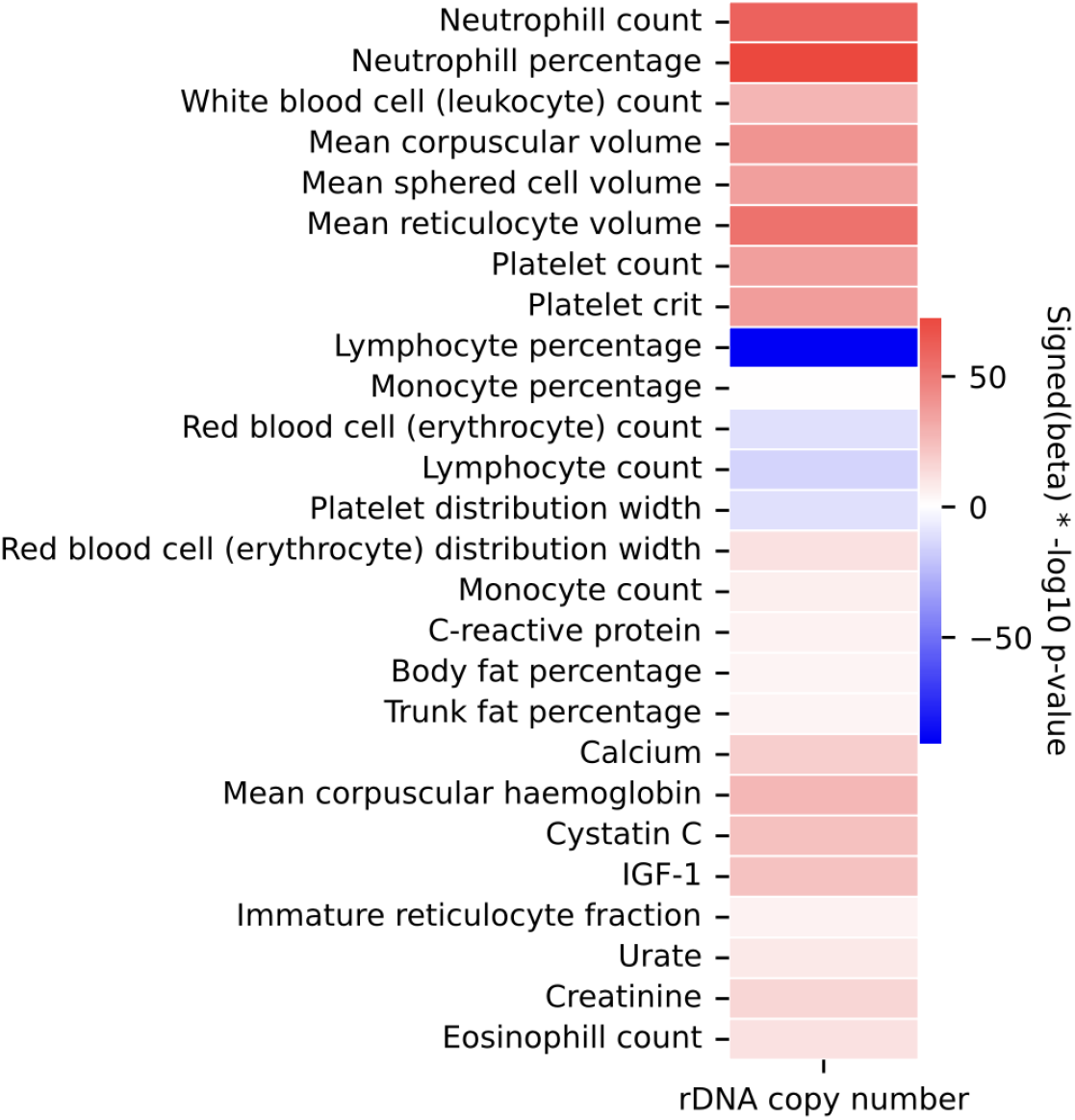
rDNA copy numbers associate with blood related traits. Heatmap of p-values for the association of rDNA copy numbers with blood traits. The heatmap color gradients indicate significance levels for each trait. The colors red and blue indicate the sign of the linear regression beta where red indicates positive association with the trait and blue for negative association.

**Figure S6:**
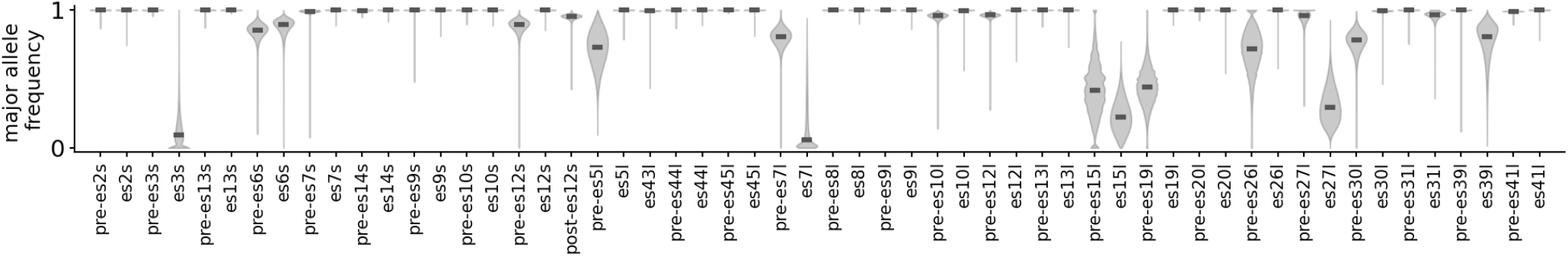
Major allele frequency of ES and non-ES regions of the 18S and 28S. Distribution of the frequencies of the most abundant allele (major allele frequency; MAF) within the UK Biobank cohort. Segments are ordered by their positions in the 18S and 28S (Table S4).

